# Association Between Nutrition Literacy and Diet Quality Among Adolescents and Young Adults in the Rural District of Mayuge, Eastern Uganda

**DOI:** 10.1101/2025.04.19.25326098

**Authors:** Thomas Buyinza, Edward Buzigi, Justine Bukenya, Mary Mbuliro, Julius Kiwanuka, Rawlance Ndejjo, David Guwatudde

**Affiliations:** Department of Epidemiology and Biostatistics, School of Public Health, Makerere University, Kampala, Uganda; Department of Community Health and Behavioural Sciences, School of Public Health, Makerere University, Kampala, Uganda; Department of Disease Control and Environmental Health, School of Public Health, Makerere University, Kampala, Uganda

**Keywords:** Adolescents, Nutrition literacy, diet quality, malnutrition, non-communicable diseases

## Abstract

**Background:** Adolescent malnutrition, including stunting, underweight, and micronutrient deficiency, is a major public health concern in Sub-Saharan Africa (SSA). While studies from the global north and Asia have shown that nutrition literacy supports healthier diets, evidence on literacy status and its role on influencing overall diet quality among adolescents and young adults (AYA) in SSA is limited. This study assessed nutrition literacy and its association with diet quality among AYA in rural Uganda.

**Method:** This was a cross-sectional study implemented as part of ARISE-NUTRINT project. Based on Nutbeam’s model of health literacy, the study was conducted among 1206 AYA aged 10–24 years in Mayuge district, Eastern Uganda, selected through stratified random sampling. Using structured questionnaire, the Global Diet Quality Score (GDQS) was adapted to estimate diet quality and Adolescent nutrition literacy scale to assess nutrition literacy status. Logistic regression models were employed to assess the association between nutrition literacy and diet quality, adjusting for socio-demographic characteristics.

**Results:** Among 1206 respondents (51.1% female), 85.9% were still in school, over 62% were from low social economic status households, and only 14% used mobile phones. Low nutrition literacy (49%) was prevalent, many unfamiliar with balanced diet or ignoring dietary advice, although 62% were willing to promote healthy eating. Overall, 12.6% had poor-diet quality based on GDQS, marked by frequent refined grain consumption and low fruits/vegetable intake. Having low nutrition literacy was associated with close to five-fold increase in poor diets (adjusted OR = 4.71, 95% CI: 2.19–10.16) while mobile phone use was associated with better diet quality by 56% (aOR = 0.44, 95% CI: 0.27–0.74).

**Conclusion:** Low nutrition literacy is a significant burden among AYA, and is strongly associated with suboptimal diet quality. Implement targeted interventions for improving nutrition literacy can enhance diet quality among AYA in the study area.

## Background

Malnutrition is one of the key global health problems among all age groups including adolescents and young adults (AYA), with more than 50% of (girls) reported to experience inadequate intake of key micronutrients such as iron, folate, zinc, calcium, and vitamins (1). In Sub-Saharan Africa (SSA), the prevalence of underweight among AYA ranged from 2.4% to 14.8%, stunting from 5.7% to 34.9%, and overweight (including obesity) from 9.2% to 11.4% in the period 2022 to 2023 (2, 3). In 2020, over 45% of adolescents in Eastern Uganda consumed diets with low-diversity, characterized by high levels of unhealthy fats, salt, and refined carbohydrates, while lacking adequate fruits and vegetables (4). These unhealthy dietary patterns put adolescents at risk of malnutrition and associated non-communicable diseases (NCDs) including anaemia, type 2 diabetes mellitus (T2DM), and cardiovascular diseases (CVDs) (5, 6).

To mitigate the risk of the triple burden of malnutrition, the existence of under-nutrition, micronutrient deficiency and overweight/obesity and associated NCDs), adolescents need to adopt and maintain high-quality diets. However, studies conducted in the SSA countries of Burkina Faso, Ethiopia, and Tanzania indicated that adolescents often struggle to adopt healthy eating lifestyles, gravitating instead toward poor-quality diets (7, 8). This could in part, be attributed to their low nutritional literacy (9, 10), besides other factors such as limitation in accessing nutrition food options (11, 12).

Adapted from Nutbeam’s tripartite model of health literacy (13), nutrition literacy refers to the ability of an individual to access, process, understand, and use basic nutrition information to make appropriate dietary or nutrition decisions (14). Nutrition literacy has three dimensions including: Functional nutrition literacy (FNL), interactive nutrition literacy (INL), and critical nutrition literacy (CNL). FNL refers to having the basic ability to read and/or comprehend dietary or nutrition labels, guidelines, and messages, while INL entails the possession of cognitive and interpersonal skills needed to communicate and engage with health professionals, peers, and others to manage personal dietary needs. On the other hand, CNL is the capacity to critically assess nutrition information, question food marketing, and advocate for improved dietary environments (participate in action to address barriers to healthy dietary intake) (14, 15).

Acquiring adequate Nutrition literacy during adolescence contributes to the development of food skills, enhances self-efficacy for healthy eating, and promotes the adoption of lifelong healthy dietary patterns (9, 10, 16). Empirical findings from the global north and other regions outside SSA, including Turkey (17, 18), Iran (19), Palestine (20), and the United States (10) has linked low nutrition literacy to unhealthy dietary choices among adolescents and adults. However, there is limited information on Nutrition literacy levels and their influence on diet quality among adolescents in the SSA region yet most of these countries report unhealthy dietary practices amidst high levels of poverty and an overreliance on subsistence agriculture. In the districts of Iganga and Mayuge in Uganda, previous research has reported unhealthy dietary patterns characterized by low dietary diversity among adolescents, (4). This study assessed Nutrition literacy status and its association with diet quality, to inform design of targeted nutrition education intervention for improving literacy and diets among adolescents and young adults in rural Eastern Uganda.

## Methods

### Study design and setting

This was a population-based cross-sectional study conducted as part of the ARISE-NUTRINT (Africa Research, Implementation Science, and Education ─ Reducing Nutrition-related NCDs in adolescence and youth: Interventions and policies to boost nutrition fluency and diet quality in Africa) initiative. Through longitudinal population-based surveys, the ARISE-NUTRINT seeks to enhance understanding of dietary and physical activity-related risks for NCDs among AYA in seven SSA countries of Uganda, Tanzania, Burkina Faso, South Africa, Ethiopia, Nigeria and Ghana, with partners from Europe and North America (21, 22). This paper focuses on the first wave (the needs assessment) of the ARISE-NUTRINT surveys for the Uganda site, implemented within the Iganga-Mayuge Health and Demographic Surveillance Site (HDSS). The Iganga-Mayuge HDSS covers three predominantly rural districts of Iganga, Mayuge and Bugweri. However, wave 1 survey for this study was conducted only in Mayuge. Under surveillance since 2005, this HDSS has 18,634 households and 120,000 residents with AYA comprising 27% of the population (23).

### Study population

The study population were AYA aged 10-24 years, sampled from the Iganga-Mayuge HDSS database with 40,000 AYA residing in Mayuge, Iganga and Bugweri districts (23). All AYA within the database and residing in Mayuge district were eligible for inclusion except those with chronic illnesses which could prevent them from adequate participation, for example those unable to speak, hear or cognitively impaired.

#### Sample size estimation

Anemia was selected as the key outcome for estimating sample because this condition was among the top five causes of morbidity in both male and female adolescents in SSA with its prevalence ranging from 20% to 60% (24, 25). Drawing from previous ARISE Network studies (22, 26, 27), it was assumed that enrolling at least 1,000 participants per country would be sufficient to detect meaningful differences in anemia rates. To account for an expected 20% loss to follow-up (25, 28, 29), the target sample size was increased to 1,200 adolescents. This sample size would ensure 80% statistical power to detect at least a 10% absolute difference in anemia prevalence between any groups compared. The power calculation was done assuming a baseline prevalence of 50%, a conservative approach because it provides the highest variability. Therefore, the design would still have adequate power across different prevalence levels within the 20–60% range

#### Sampling procedures

From the Iganga-Mayuge HDSS database, eligible AYA were randomly selected using household-based simple random sampling within each age-sex stratum (10–14, 15–19, and 20– 24 years; male and female), applying Probability Proportional to Size allocation to each stratum. In households with more than one eligible AYA, only one was randomly selected to participate using a within-household randomization procedure, to minimize intra-household correlation. A list of sampled AYA was generated, including locator information for their households. Those selected were then approached in their households using this locator information, with support from community guides.

#### Conceptual framework

Analysis for this study was premised on a framework adapted from Nutbeam’s Model of Health Literacy. In this model Nutrition literacy is conceptualized as a sub-component of health literacy, which, in turn, is a component of the broader general literacy (30-32). Nutrition literacy is further categorized into three dimension: FNL, INL, and CNL Nutbeam (32) and previously in Uganda, this three-level categorization was applied to assess Nutrition literacy status of adolescent students in Kampala city (33). While Ndahura (33) used a total of 29 attitude statements, this study refined this approach by adapting 22 statements to measure Nutrition literacy among the AYA in rural Mayuge District in Uganda, considering contextual differences, such as lower formal education. access, socioeconomic status as well as reducing time and cost during data collection to meet feasibility requirements. Nutrition literacy was hypothesized as a key factor influencing food choices and eating behaviors (10, 17, 19, 20). This highlights its role in shaping overall diet quality among AYA, presented in **supplementary information: figure 1**.

### Data collection and Measurements of study variables

The current study utilized a longitudinal survey questionnaire for the ARISE-NUTRINT initiative. This tool was translated into *Lusoga*, the major local language spoken in Mayuge District, and uploaded onto Open Data Kit for electronic data collection. Trained research assistants conducted one-on-one, face-to-face interviews between April and May 2024. Data were subjected to iterative cross-checks and reviews for completeness, and in cases of missing information, participants were contacted for follow-up to address the gaps.

#### Diet quality (outcome variable)

The Global Diet Quality Score (GDQS), a food-based index structured as a food-frequency questionnaire, was adapted to measure diet quality (34, 35). This scale was previously validated in 10 SSA countries (including Uganda), India, China and the United States of America (35, 36). Unlike the Dietary Diversity Score which only assesses the risk of micronutrient deficiency (37, 38), the GDQS assess overall diet quality in relation to both nutrient adequacy and risk of NCDs, for instance, the score reflect risks for the triple burden of malnutrition: undernutrition (wasting, underweight and stunting), micronutrient deficiency (anemia, increased infection rates), and overnutrition (overweight, obesity and associated NCDs like CVDs and T2DM) (35, 36). Previously, higher GDQS scores were reported to be associated with improved nutrient adequacy, better mid-upper arm circumference, and reduced anemia risk among adolescents in SSA, including Uganda(36). Although the GDQS was validated using food amounts/servings (grams/day) for cross-country and temporal comparisons, the present study adapted this index to measure diet intake using frequency rather than quantity, as it was done in past studies in SSA due to the lack of standardized serving sizes in rural, low-income settings (27, 39).

Basing on nutrient adequacy and nutrient requirements for AYA, food consumed was classified into 25 food groups, split into 17 healthy and 8 unhealthy groups. The healthy category included dark green vegetables, legumes and nuts, and seeds, fresh fruits, cruciferous vegetables, deep orange vegetables, deep orange tubers, and other vegetables, lean meat, white meat, eggs, low and high-fat milk, whole grains, boiled snacks, matooke, root, and stem tubers). On the other hand, unhealthy food groups included unprocessed red meat, deep-fried foods, snacks high in salt or oils/fat, refined grains, processed meat, juice, white roots and tubers, sugar-sweetened beverages and sweets and ice cream. High-fat milk and dairy products were classified as healthy food groups at all levels of consumption. This was an adaptation of the original GDQS, which categorized high consumption (≥3 times per day) of high-fat dairy as unhealthy and scored it negatively (35). In the context of the rural, low-income setting of the current study, both low- and high-fat dairy were considered healthy, given the important role dairy plays as a nutrient source for the adolescents and young adults (27).

Participants were asked how often, on average, they had consumed each food group, over the past month and responses recorded as 0-1/week, 2-3/week or 4+/week were used to score diet quality according to food groups consumed more or less frequently. Following the GDQS scoring guidelines, lower points were assigned for less frequent consumption of healthy food groups, while higher points were assigned for less frequent consumption of unhealthy food groups (35). For example, from the healthy foods; dark green vegetables, legumes and nuts, and seeds, fresh fruits: scored as 0–1 (0 points), 2–3 (2 points), ≥4 (4 points); and from the unhealthy groups; deep-fried foods, snacks high in salt or oils/fat, refined grains, processed meat, juice, white roots and tubers. Sugar-sweetened beverages and sweets: 0–1 (2 points), 2– 3 (1 point), ≥4 (0 points). The detailed scoring plan used is presented as **Supplementary Information: Table 1**.

For each participant, diet quality scores were summed to obtain an overall GDQS (0 to 49) with higher scores indicating better diet quality. Participants with a GDQS <15 were classified as having poor-quality diets (associated with increased risk of nutrient inadequacy and higher risk of diet-related NCDs) (35). Those with scores ≥15 were classified as not having poor-quality diets (associated with reduced risk of nutrient inadequacy and lower risk of diet-related NCDs) (35). This binary categorization of diet quality was considered because it matches with population-level nutrition interventions aimed at improving diets from poor quality to healthy (not poor) and it simplifies interpretation, and facilitate clear messaging for nutrition policymakers and program implementers (40, 41).

#### Nutrition literacy (major independent variable)

Nutrition literacy (with its three dimensions) was measured using the adolescent Nutrition Literacy Scale (ANLS) with 22 attitude statements, each answered on a 5 point-Likert scale (33). The FNL dimension has seven items: with possible scores ranging from 0-7; INL has six items with possible scores ranging from 0-6 and CNL has nine items with possible scores ranging from 0-9 (33). Twelve statements of the ANLS were intentionally reversed to trigger respondents’ attention, reduce automatic responses, and promote thoughtful engagement through balanced framing. The reversing also help identify inconsistencies, enhancing scale reliability and measurement precision (33). Direct statements for instance “I am familiar with the concept of a balanced diet” were scored 0 for strongly disagree, disagree, and neither agree nor disagree, and 1 for agree and strongly agree. Reversed statements for example “I find it difficult to know how I should change my diet when I get dietary advice from the doctor, nurse, or the like” were scored 1 for strongly disagree and disagree, and 0 for neither agree nor disagree, agree, and strongly agree as shown in the **supplementary information: table 2**.

Since Likert scales measure ordinal data with unequal intervals between responses, scoring them as continuous numbers would lead to misinterpretation (42). To address this, scores were converted into binary responses, treating "neither agree nor disagree" as incorrect (scored 0) (33). Median was used to summarize FNL, INL, and CNL subscales instead of mean, as it respects ranking of the Likert scores without assuming equal intervals (42). Overall nutrition literacy was computed by summing subscale scores, with possible totals ranging from 0 to 22 (higher scores indicating higher nutrition literacy). Due to limited literature, total scores were categorized into low (≤7), moderate (8–14), and high (≥15) basing on tertiles, adapting previous studies which used data-driven cut-offs in the absence of validated thresholds (33, 43).

#### Confounding variables

In addition to nutrition literacy, potential confounders such as individual and parental demographics like socio-economic characteristics (SES), household food security status, phone ownership, and social media use were included in the questionnaire. Social media use was assessed by asking respondents if they had an account on any of Facebook, WhatsApp, TikTok, or Telegram (yes/no), and those with at least one account on any of these channels were categorized as using social media. Besides, individual ownership of a phone or using a parent’s, neighbour’s or friend’s phones was reported as phone use. Household food security status was assessed using a question “in the past 30 days, was there ever no food to eat of any kind in your house because of lack of resources to get food?” with Yes/No as the response options. The MacArthur Scale of Social Status was adopted to measure SES. This scale depicts social status as a 10-rung ladder, where individuals perceive social status within their community and society as a whole (44). Participants were asked to state the rung on the ladder on which they feel their family stood at the moment during data collection. On a scale of 0-10, higher scores indicated better SES within their community or society at large (more resources, respect, or influence), where 1-3 was categorized as low, 4-6 as mid, and 7-10 as high SES (44).

### Statistical analyses

Data analysis was conducted using STATA version 15.0. To characterize the AYA included in the study, we used frequencies and percentages for ordinal and nominal data and the mean with standard deviation or median plus inter-quartile range (IQR) for the Likert scores based on the normality distribution for the transformed score variables. The Cronbach’s alpha coefficient for the Nutrition literacy and diet quality scales were 0.70 and 0.75 respectively, indicating reliability reliable scale. Considering diet quality as a binary outcome categorized into poor quality (coded as yes or 1 and not poor (coded as no or 0), logistic regression was used to determine the crude associations between diet quality and nutrition literacy. Separate bivariate logistic regression models were run between the binarized outcome and each of the other independent variables including age, sex, parental education level, phone use and social media use. Multicollinearity was assessed using the Pearson correlation coefficient (r), retaining only variables with a stronger biological relevance or better model fit if r > 0.4. The multivariable logistic regression model was built using backward stepwise process, where the least significant covariate based on p-value was removed at each step. Statistical significance P < 5% and adjusted odds ration were used to determine the magnitude and direction of the association between diet quality and nutrition literacy. Afterwards, non-significant covariates were individually adjusted in the multivariable model to determine their adjusted odds ratios.

### Ethical considerations

This study, part of the ARISE-NUTRINT project for the Uganda site, was approved by the Research and Ethics Committee at the School of Public Health, Makerere University (Reference number: SPH-2023-460). Written informed consent was obtained from parents/guardians of adolescents (10-17 years), followed by adolescent informed assent. Adults (18+ years) provided written informed consent. Interviews were conducted privately to ensure confidentiality. Data collected had no personal identifiers and was securely stored on password-protected computers and servers, restricting access to authorized study team members only.

## RESULTS

### Socio-demographic characteristics

**Table 1** summarizes the socio-demographic characteristics of the respondents. A total of 1206 AYA participated in the study, 51.1% being males. The majority of respondents (76.4.8%) were aged 10 to 14 years and over 85% were in school. Approximately three-quarters (74.8%) of the respondents came from moderately food-insecure Households while none resided in severely food-insecure Households. The SES was moderate for most respondents (62.4%) with only 4.6% perceiving to be in high SES category. The proportion of phone use was 14% and only 3.3% of the respondents reported using social media. Regarding the education status of the parents, only 24% had fathers or male guardians who had attained secondary education or higher.

**Table 1:** Socio-demographic characteristics of the respondents (n=1206)

| Variable | Frequency (n) | Percentage (%) |
| --- | --- | --- |
| **Age group** |  |  |
| 10-14 | 922 | 76.4 |
| 15-19 | 190 | 15.8 |
| 20-24 | 94 | 7.8 |
| **Sex** |  |  |
| Female | 616 | 51.1 |
| Male | 590 | 48.9 |
| **Currently in school** |  |  |
| No | 170 | 14.1 |
| Yes | 1036 | 85.9 |
| **Household Food insecurity** |  |  |
| No | 939 | 77.9 |
| Yes | 267 | 22.1 |
| **Socio-economic status** |  |  |
| Low | 753 | 62.4 |
| Moderate | 397 | 33.0 |
| High | 56 | 4.6 |
| **Mobile phone use** |  |  |
| No | 1037 | 86.0 |
| Yes | 169 | 14.0 |
| **Social media use** |  |  |
| No | 1166 | 96.7 |
| Yes | 40 | 3.3 |
| **Father's education level** |  |  |
| Below Primary | 200 | 17.5 |
| Primary | 670 | 58.5 |
| Secondary and above | 275 | 24.0 |
| **Mother's education level** |  |  |
| Below Primary | 222 | 18.7 |
| Primary | 763 | 64.1 |
| Secondary and above | 205 | 17.2 |

### Diet quality

Over 29.2% (352/1206) consumed refined grains and flour products more than four times per week (wk), on average, in the month preceding data collection. Additionally, 19.8%, 32.1%, and 27.9% consumed purchased deep-fried foods, Sugar-sweetened beverages, and sweets/ice cream 2–3 times per week, respectively. Less than a quarter (23.6%) consumed leafy vegetables/fruits 4+ times per week. The mean GDQS (±SD) was 21.5 (±4.8) (range 11-37.5); and overall, 12.6% (152/1206) of the participants had GDQS scores falling into the poor-quality category as shown in Table 2.

**Table 2:** Proportion of responses across the food consumption frequency on the GDQS (n=1206)

| Question | How often, on average, have you consumed each food group, over the past month? | Percentage response. |  |  |
| --- | --- | --- | --- | --- |
|  | Food group | 0-1/wk | 2-3/wk | 4+/wk |
| Healthy food groups | Legumes & derived products (e.g., tofu, soymilk) | 53 | 41.5 | 5.5 |
|  | Deep orange vegetables (except roots & tubers) | 54.2 | 43.2 | 2.6 |
|  | Dark green leafy vegetables<br>(Examples of local dark green leafy vegetables) | 24.1 | 52.4 | 23.6 |
|  | Cruciferous vegetables (lettuce, cabbage, cauliflower, etc.) | 46.4 | 46.5 | 7.1 |
|  | Other vegetables (except roots & tubers) | 52.4 | 41.1 | 6.5 |
|  | Citrus fruits (orange, lime, lemon, etc.) | 55.6 | 39.1 | 5.2 |
|  | Deep orange fruits like passion fruits, mangoes, watermelon, pawpaw | 60.7 | 36.4 | 2.9 |
|  | Other fruits like bananas, avocado, jackfruits, tamarind | 49.9 | 41.1 | 9.0 |
|  | Deep orange tubers like carrots, orange-flesh sweet potatoes | 60.2 | 29.8 | 10.0 |
|  | Nuts and seeds (incl. nut butter) | 30.0 | 60.9 | 9.1 |
|  | Liquid vegetable oils | 15.6 | 46.5 | 37.9 |
|  | Poultry and other bird meat | 71.9 | 26.7 | 1.4 |
|  | Fish | 56.4 | 40.3 | 3.3 |
|  | Whole (i.e., brown) grains & flour products | 48.3 | 41.0 | 10.7 |
|  | Eggs | 60.5 | 35.8 | 3.7 |
|  | Low-fat milk & dairy products | 72.4 | 24.0 | 3.6 |
|  | High-fat milk & dairy products (except butter) | 68.9 | 27.7 | 3.4 |
| Unhealthy food groups | Refined (i.e., white) grains & flour products | 27.0 | 43.8 | 29.2 |
|  | Unprocessed red meat (and organs) | 58.4 | 41.6 | 0 |
|  | Processed meat (e.g., salted, cured, smoked) | 80.9 | 17.7 | 1.4 |
|  | White roots & tubers (flours, e.g., potato flour) | 42.4 | 39.8 | 17.8 |
|  | Sweets and ice cream | 66.3 | 27.9 | 5.8 |
|  | Sugar-sweetened beverages | 64.3 | 32.1 | 3.6 |
|  | Purchased deep-fried foods | 78.3 | 19.8 | 1.9 |
|  | Fruit juice (with added sugar) | 70.0 | 27.9 | 2.1 |
|  | Food group | 0-1/wk | 2-3/wk | 4+/wk |
|  | **Overall diet quality, based on GDQS** |  |  |  |
| | Mean (±SD) | 21.5 (±4.8) | | |
|  | **Poor-quality** |  |  |  |
|  | Yes (%) | 12.6 |  |  |
|  | No (%) | 87.4 |  |  |

### Nutrition literacy status

Table 3 summarizes the proportion of responses across Likert scale for each of the indicators of Nutrition literacy. None of the respondents strongly agreed that they were familiar with the recommendation for daily intake of fruits and vegetables for their age and none found it easy to understand when they read or heard information about nutrition, food, or diet. Concerning the “balanced diet”, only 1.8% (22/1206) strongly agreed that they were familiar with this concept and only 1.4% discussed diet with friends and family. While the majority (52%) agreed that they found it easy to change their diet after receiving professional advice, only 22.6% had altered eating habits based on dietary information gathered. However, more than a quarter (29.9%) agreed that they try to influence others like family and friends to eat healthy food and over 62% were willing to take an active role in promoting a healthier diet in their respective communities. The median FNL score was 1, with an interquartile range (IQR) of 0–3, and only 0.17% achieved the maximum possible score of 7 (scale: 0–7). For the INL dimension, the median score was 2, IQR: 1–3 while for CNL, the median score was 4, IQR: 3–5 and only 0.25% had a maximum score of 9 (scale: 0–9). Overall, close to half (49%) of the respondents demonstrated low nutrition literacy and only 3.7%e scored in the high literacy category.

**Table 3:**
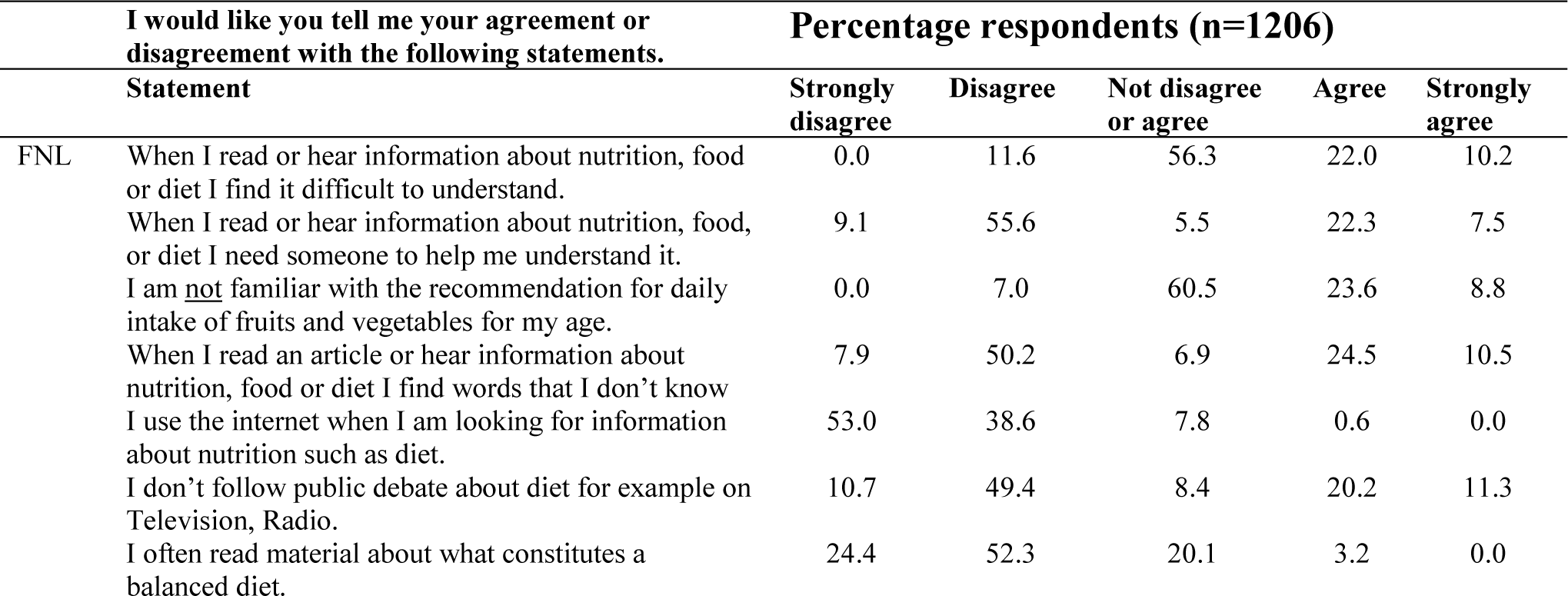
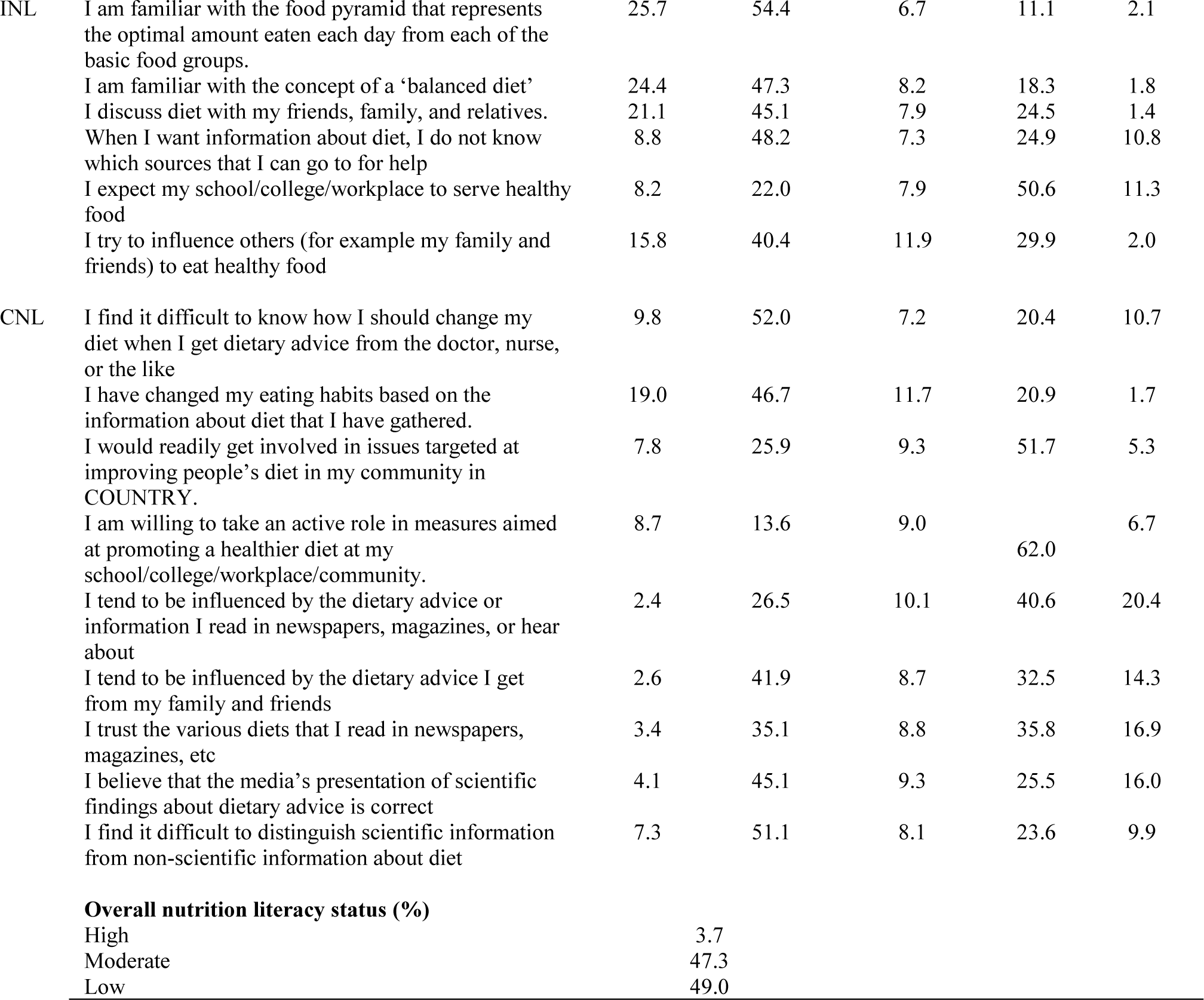
Proportion of responses across Likert scale options on the ANLS.

### Association between nutrition literacy and diet quality

At bivariable analysis, low and moderate nutrition literacy status were significantly associated with poor diet quality (crude OR=3.95, 95% CI: 1.96-7.97) and (crude OR=2.36, 95% CI: 1.19-4.67) respectively. Besides, low SES (crude OR=2.95. 95%CI: 1.51-5.75) and phone use (crude OR=0.59, 95%CI: 0.38-0.91) also had significant crude associations with poor diet quality. At multivariable analysis, mother’s education variable was dropped due to a strong correlation with the father’s ’s education (r>0.4) and similarly, currently in school was dropped due to strong negative correlation with age (r>-0.4). When qualifying variables were adjusted for in a multivariable logistic regression model, respondents with low and moderate nutrition literacy were 4.71 and 2.73 times more likely to have poor-diet quality (adjusted OR = 4.71, 95% CI: 2.19-10.16) and moderate (adjusted OR=2.73, 95% CI: 1.31-5.70) compared to those with high nutrition literacy. In addition to literacy, having low SES was associated with 2.74 times higher likelihood of poor diet quality (adjusted OR = 2.74, 95% CI: 1.28-5.86); while those who used mobile phones were 56% less likely to have poor diet quality (adjusted OR = 0.44, 95% CI: 0.27–0.74) as shown in Table 4.

**Table 4:** Association between nutrition literacy and diet quality (n=1206)

| Variable | Poor- Quality Diet |  | Crude odds ratios | Adjusted odds ratios |
| --- | --- | --- | --- | --- |
|  | Yes n (%) | No n (%) | (95% CI) | (95% CI) |
| **Nutrition literacy** |  |  |  |  |
| High | 13 (28.9) | 32(71.1) | 1 | 1 |
| Moderate | 84 (14.7) | 487 (5.3) | **2.36 (1.19-4.67) \*** | **2.73 (1.31-5.70) \*\*** |
| Low | 55 (9.3) | 535(90.7) | **3.95 (1.96-7.97) \*\*\*** | **4.71 (2.19-10.16) \*\*\*** |
| 10-14 | 114 (12.4) | 808 (87.6) | 1 | 1 |
| 15-19 | 27 (14.2) | 163 (85.8) | 0.85 (0.54-1.34) | 1.11 (0.67-1.82) |
| 20-24 | 11 (11.7) | 83 (88.3) | 1.06 (0.55-2.06) | 1.98 (0.89-4.40) |
| **Sex** |  |  |  |  |
| Male | 84 (14.2) | 5060 (85.8) | 1 | 1 |
| Female | 68 (11.0) | 548 (89.0) | 1.34 (0.95-1.88) | 1.23 (0.86-1.77) |
| **Currently in school** |  |  |  |  |
| No | 19 (11.2) | 151 (88.8) | 1 |  |
| Yes | 133 (12.8) | 903 (87.2) | 0.85 (0.51-1.42) |  |
| **Household food insecurity** |  |  |  |  |
| No | 117 (12.5) | 822 (87.5) | 1 | 1 |
| Yes | 35 (13.1) | 232 (86.9) | 0.94 (0.63-1.41) | 0.85 (0.55-1.32) |
| **Socio-economic status** |  |  |  |  |
| High | 13 (23.2) | 43 (76.8) | 1 | 1 |
| Moderate | 69 (17.4) | 328 (82.6) | 1.44 (0.73-2.82) | 1.23 (0.58-2.6) |
| Low | 70 (9.3) | 683(90.7) | **2.95 (1.51-5.75) \*\*** | **2.74 (1.28-5.86) \*\*** |
| **Father's education level** |  |  |  |  |
| Below primary | 29 (14.5) | 171 (85.5) | 1 | 1 |
| Primary | 86 (12.8) | 584 (87.2) | 1.15 (0.73-1.81) | 1.19 (0.74-1.93) |
| Secondary and above | 27 (9.8) | 248 (90.2) | 1.55 (0.89-2.72)) | 1.74 (0.97-3.10) |
| **Mother's education level** |  |  |  |  |
| Below Primary | 23 (10.4) | 199 (89.6) | 1 |  |
| Primary | 105 (13.8) | 658 (86.2) | 0.72 (0.45-1.17) |  |
| Secondary and above | 18 (8.8) | 187(91.2) | 1.20 (0.63-2.30) |  |
| **Mobile phone use** |  |  |  |  |
| No | 121 (11.7) | 916 (88.3) | 1 | 1 |
| Yes | 31 (18.3) | 138 (81.7) | **0.59 (0.38-0.91) \*** | **0.44 (0.27-0.74) \*\*** |
| **Social media use** |  |  |  |  |
| No | 148 (12.7) | 1018 (87.3) | 1 | 1 |
| Yes | 4 (10.0) | 36 (90.0) | 1.31 (0.46-3.73) | 2.05 (0.66-6.37) |

p<0.001 = \*\*\*, p<0.01 = \*\*, p<0.05 = \*

## DISCUSSION

This study assessed nutrition literacy status and its association with diet quality among AYA in the rural settings of Mayuge districts in Uganda. The majority of the respondents had low nutrition literacy, where none strongly agreed that they were familiar with the recommendation for daily intake of fruits and vegetables for their age and only a few had altered eating habits based on dietary information obtained from health workers. Besides, the majority frequently consumed refined grains and flour products (more than four times per week), and less than a quarter consumed green leafy vegetables 4+times per week. Having low nutrition literacy was strongly associated with poor diet quality and other significant correlates of poor diet quality included having low SES, and not using mobile phones.

In the present study, only 3.7% of the respondents had high nutrition literacy and the majority (49%) demonstrated low literacy status. Over 80% of respondents were unfamiliar with the food pyramid and the majority had difficulties in comprehending diet and nutrition information. These findings are consistent with a study in Turkey which found suboptimal levels of nutrition literacy among early adolescents (45). Contrary to our findings, the majority of the adolescents in urban Uganda (Kampala city) exhibited moderate levels of nutrition literacy, with basic skills required to comprehend and follow nutrition messages (33). Adolescents in the rural settings for our study may have limited access to nutrition and health information and lower general literacy, leading to low nutrition literacy compared to their urban counterparts in the Kampala study, (33). Related past studies found higher levels of FNL, INL, CNL, and overall nutrition literacy among adolescents in urban settings than those who resided in rural areas (17, 46). Individuals with poor nutrition literacy levels have limited basic nutritional knowledge and cannot critically analyze food and nutrition information to make their dietary decisions (47). Therefore, our study findings underscore the urgent need for tailored nutrition education interventions to improve nutrition literacy among’ adolescents in rural Uganda.

The majority of respondents exhibited poor diet quality, with 71% consuming refined grains and 32.8% drinking sugar-sweetened beverages more than four times per week. Only 23.3% consumed dark green leafy vegetables at least 2-3 times per week. These findings align with a study in Iganga and Mayuge, Uganda, where 45% of adolescents had low dietary diversity, marked by high consumption of fats and oils but insufficient fruits and vegetables (4). Consistent patterns were observed in urban Uganda, where 50% and 76% of adolescents reported inadequate fruit and vegetable consumption, respectively (48). Similar unhealthy dietary patterns have been reported in other sub-Saharan African countries, including Ethiopia (49), Ghana (11), and Ethiopia (50). Several factors may explain the low vegetable intake in the present study, including negative sensory attributes of vegetables, such as bitter, bland, or pungent flavors and undesirable textures like mushiness (51). Cultural dietary norms (52), and limited knowledge about the nutritional benefits of fruits and vegetables (53) also contribute while the high consumption of sugar-sweetened beverages may be attributed to the proliferation of carbonated drinks due to globalization (11). The poor-quality diets indicate a high risk of micronutrient deficiency, undernutrition, overnutrition, and diet-related NCDs (35, 54, 55).

Our study identified a significant positive association between Nutrition literacy and diet quality levels, among AYA in rural Eastern Uganda, with poor Nutrition literacy predicting a threefold increase in the likelihood of having poor-quality diets. This is consistent with past studies in low-middle income countries (LMICs) like Turkey (17), Iran (56, 57) and Taiwan (58), and in high-income countries like the USA ((10), where higher nutrition literacy levels correlated with healthier eating habits among adolescents. For instance, in Turkey, adolescents with higher nutrition literacy consumed more fruits and vegetables, while those with lower levels had frequent intake of unhealthy snacks (17, 59). This emphasizes the crucial role of nutrition literacy in enabling informed dietary choices and facilitating better-quality diets. In a similar context, earlier evidence suggested that food literacy may play a role in shaping youth eating behaviors (16, 60), and a global systematic review reported that adolescents with high food literacy levels followed healthier dietary practices (61).

Other than nutrition literacy, several socio-demographic factors also had a significant influence on the diet quality among AYA in rural Uganda. Low SES was significantly associated with poor diet quality, consistent with findings of studies in Ethiopia (50), South Africa (62), and Cameroon (63). This is plausible due to the varying ability to access diverse and nutritious foods across SES levels based on income status and or household food production diversity (64). Although only 14% used mobile phones, such respondents were less likely to have poor-quality diets, reflecting the potential of leveraging technology to disseminate healthy diet awareness and nutrition education messages to enhance adolescents’ self-efficacy and ultimately improve diet quality and nutrition outcomes (Kulandaivelu et al., 2023). Nonetheless, it is essential to consider that not all mobile phone use is beneficial; for example, some evidence in Qatar suggests that social media engagements on phone may be linked to unhealthy dietary habits, possibly due to exposure to persuasive food advertising or peer influences (65).

### Study limitations and strengths

This study is the first to assess the link between nutrition literacy and diet quality using the Global Diet Quality Score among AYA in a rural, low-income setting. Despite its novelty, the wide confidence interval (2.19–10.16) signals uncertainty, likely due to a small sample size, measurement errors, and population variability. Self-reported data may have introduced recall and social desirability biases. Nonetheless, validated tools showed good reliability (Cronbach’s alpha: 0.70 for nutrition literacy, 0.75 for diet quality), and analyses were adjusted for parental education and household food security. The study offers early evidence that low nutrition literacy may contribute to overall poor diet quality (including increased risk of nutrient inadequacy and diet-related NCDs) among rural AYA. Future research should use larger samples and objective measures to improve precision and deepen understanding of this relationship.

### Conclusion

The majority of the study respondents exhibited low nutrition literacy and none had high levels. Profound gaps in literacy included unfamiliarity with a balanced diet and difficulty changing dietary habits despite receiving professional advice. Diet quality was significantly better among adolescents and young adults with high nutrition literacy compared to those with low literacy. Besides, respondents with moderate SES, and those who used mobile phones had better diet quality. To enhance improvement in diet quality, there is a need to design and implement nutrition education interventions tailored to adolescents in rural setting context, to improve nutrition literacy, especially for those, those from low socioeconomic status households and those with no access to mobile phones.

## Data Availability

The dataset analyzed for the present study is not publicly available due [being kept confidential for other on-going and future analysis] but can be provided by the corresponding author (TB) upon reasonable request.

## Supplementary information

**Figure. 1:**
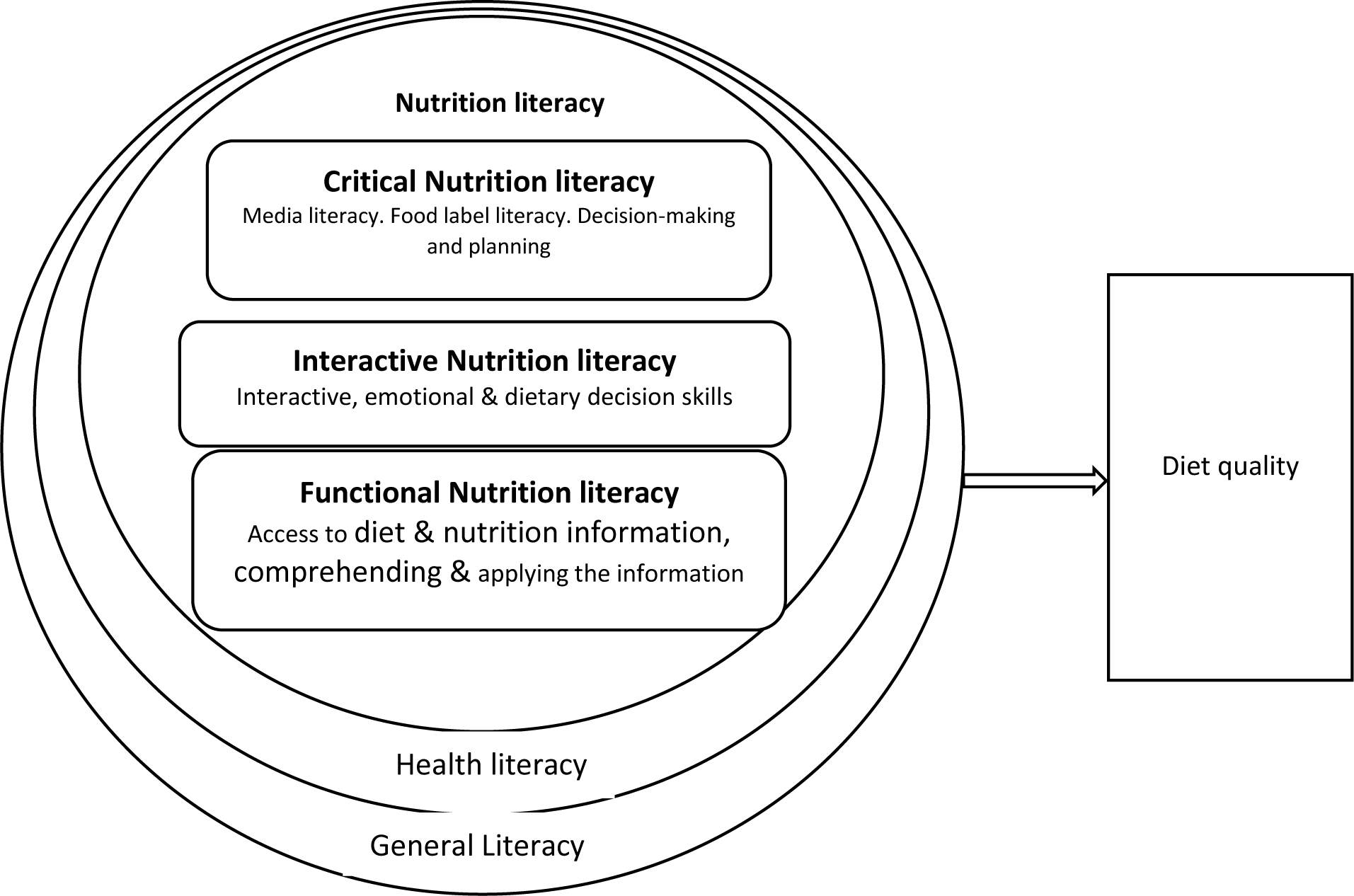
Conceptual framework illustrating the dimensions of nutrition literacy and its relationship with diet quality: adapted from the Nutbeam’s Model of Health literacy.

**Table 1:** Scores on the adolescent nutrition literacy scale, across functional (FNL), interactive (INL), and critical (CNL) subscales.

|  | I would like you tell me your agreement or disagreement with the following statements. | Strongly disagree | Disagree | Not disagree or agree | Agree | Strongly agree |
| --- | --- | --- | --- | --- | --- | --- |
| Reversed statements | When I read or hear information about nutrition, food or diet I find it difficult to understand (FNL). | 1 | 1 | 0 | 0 | 0 |
|  | I find it difficult to know how I should change my diet when I get dietary advice from the doctor, nurse, or the like (CNL). | 1 | 1 | 0 | 0 | 0 |
|  | When I read or hear information about nutrition, food, or diet I need someone to help me understand it (FNL). | 1 | 1 | 0 | 0 | 0 |
|  | I am <u>not</u> familiar with the recommendation for daily intake of fruits and vegetables for my age (FNL). | 1 | 1 | 0 | 0 | 0 |
|  | When I want information about diet, I do not know which sources that I can go to for help (INL). | 1 | 1 | 0 | 0 | 0 |
|  | I don't follow public debate about diet for example on Television, Radio (FNL). | 1 | 1 | 0 | 0 | 0 |
|  | When I read an article or hear information about nutrition, food or diet I find words that I don't know (FNL) | 1 | 1 | 0 | 0 | 0 |
|  | I tend to be influenced by the dietary advice or information I read in newspapers, magazines, or hear about (CNL). | 1 | 1 | 0 | 0 | 0 |
|  | I tend to be influenced by the dietary advice I get from my family and friends (CNL). | 1 | 1 | 0 | 0 | 0 |
|  | I trust the various diets that I read in newspapers, magazines, etc (CNL). | 1 | 1 | 0 | 0 | 0 |
|  | I believe that the media's presentation of scientific findings about dietary advice is correct (CNL). | 1 | 1 | 0 | 0 | 0 |
|  | I find it difficult to distinguish scientific information from non-scientific information about diet (CNL). | 1 | 1 | 0 | 0 | 0 |
| Direct statements | I am familiar with the food pyramid that represents the optimal amount eaten each day from each of the basic food groups (INL). | 0 | 0 | 0 | 1 | 1 |
|  | I am familiar with the concept of a ‘balanced diet’ (INL) | 0 | 0 | 0 | 1 | 1 |
|  | I use the internet when I am looking for information about nutrition such as diet (FNL). | 0 | 0 | 0 | 1 | 1 |
|  | I discuss diet with my friends, family, and relatives (INL). | 0 | 0 | 0 | 1 | 1 |
|  | I have changed my eating habits based on the information about diet that I have gathered (CNL). | 0 | 0 | 0 | 1 | 1 |
|  | I often read material about what constitutes a balanced diet (FNL). | 0 | 0 | 0 | 1 | 1 |
|  | I would readily get involved in issues targeted at improving people’s diet in my community in COUNTRY (CNL). | 0 | 0 | 0 | 1 | 1 |
|  | I am willing to take an active role in measures aimed at promoting a healthier diet at my school/college/workplace/community (CNL). | 0 | 0 | 0 | 1 | 1 |
|  | I expect my school/college/workplace to serve healthy food (INL). | 0 | 0 | 0 | 1 | 1 |
|  | I try to influence others (for example my family and friends) to eat healthy food (INL). | 0 | 0 | 0 | 1 | 1 |

**Table 2:**
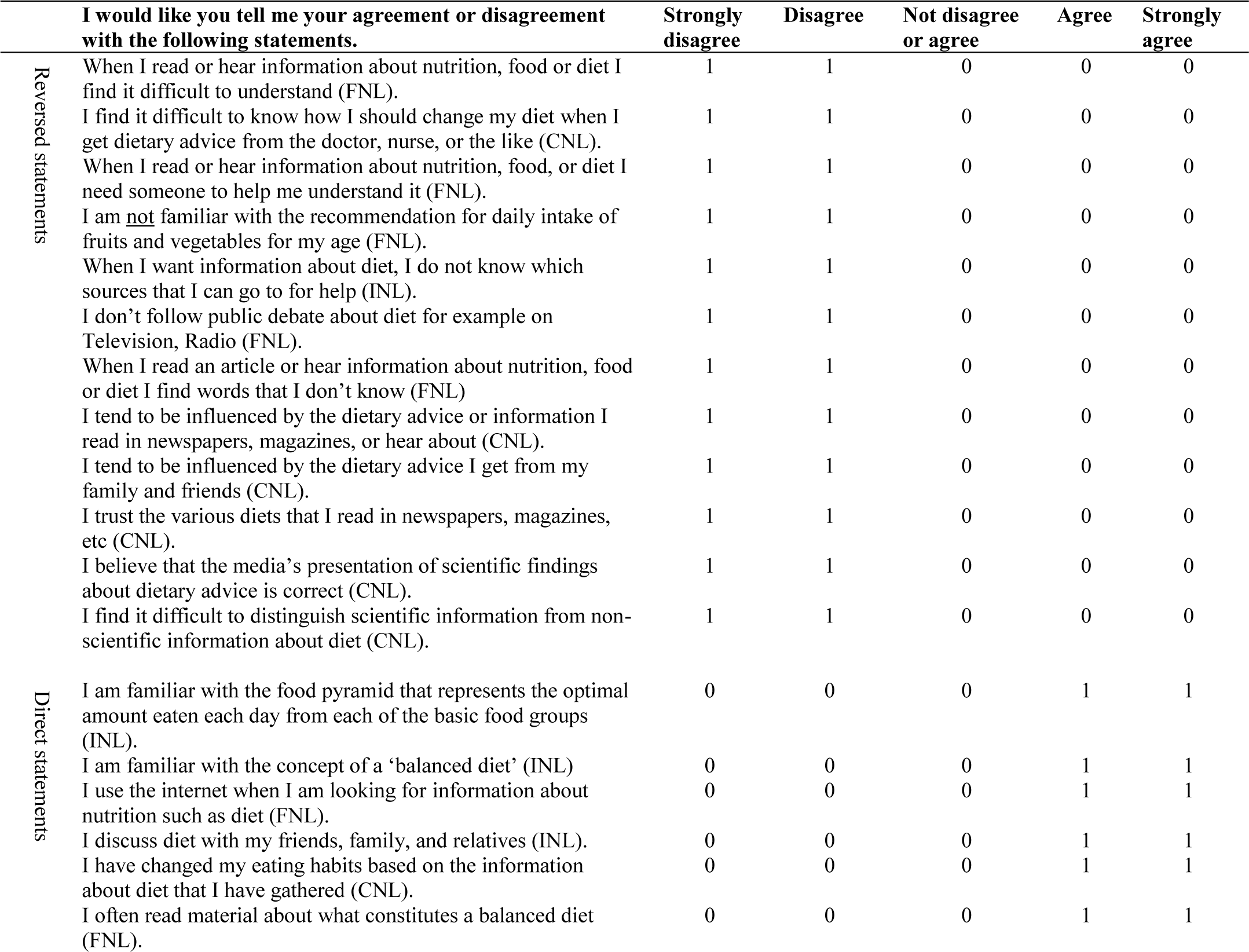
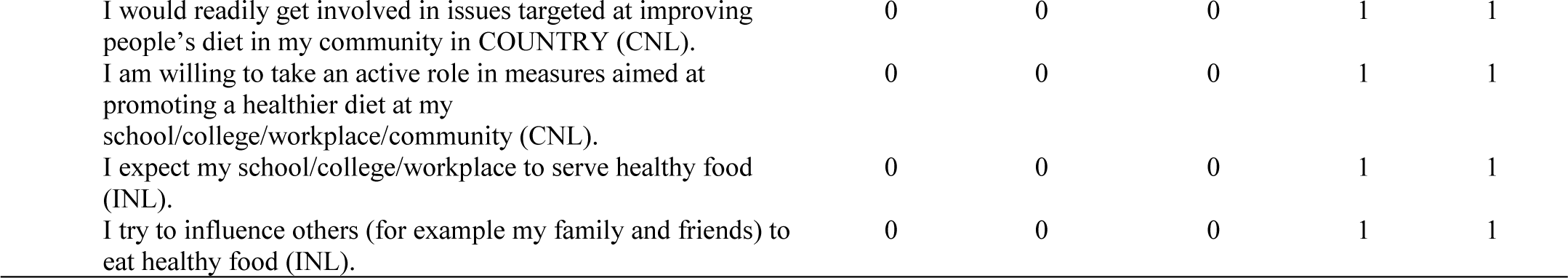
Scores on the adolescent nutrition literacy scale, across functional (FNL), interactive (INL), and critical (CNL) subscales.

|  | **I would like you tell me your agreement or disagreement with the following statements.** | **Strongly disagree** | **Disagree** | **Not disagree or agree** | **Agree** | **Strongly agree** |
| --- | --- | --- | --- | --- | --- | --- |
| Reversed statements | When I read or hear information about nutrition, food or diet I find it difficult to understand (FNL). | 1 | 1 | 0 | 0 | 0 |
|  | I find it difficult to know how I should change my diet when I get dietary advice from the doctor, nurse, or the like (CNL). | 1 | 1 | 0 | 0 | 0 |
|  | When I read or hear information about nutrition, food, or diet I need someone to help me understand it (FNL). | 1 | 1 | 0 | 0 | 0 |
|  | I am <u>not</u> familiar with the recommendation for daily intake of fruits and vegetables for my age (FNL). | 1 | 1 | 0 | 0 | 0 |
|  | When I want information about diet, I do not know which sources that I can go to for help (INL). | 1 | 1 | 0 | 0 | 0 |
|  | I don’t follow public debate about diet for example on Television, Radio (FNL). | 1 | 1 | 0 | 0 | 0 |
|  | When I read an article or hear information about nutrition, food or diet I find words that I don’t know (FNL) | 1 | 1 | 0 | 0 | 0 |
|  | I tend to be influenced by the dietary advice or information I read in newspapers, magazines, or hear about (CNL). | 1 | 1 | 0 | 0 | 0 |
|  | I tend to be influenced by the dietary advice I get from my family and friends (CNL). | 1 | 1 | 0 | 0 | 0 |
|  | I trust the various diets that I read in newspapers, magazines, etc (CNL). | 1 | 1 | 0 | 0 | 0 |
|  | I believe that the media’s presentation of scientific findings about dietary advice is correct (CNL). | 1 | 1 | 0 | 0 | 0 |
|  | I find it difficult to distinguish scientific information from non-scientific information about diet (CNL). | 1 | 1 | 0 | 0 | 0 |
| Direct statements | I am familiar with the food pyramid that represents the optimal amount eaten each day from each of the basic food groups (INL). | 0 | 0 | 0 | 1 | 1 |
|  | I am familiar with the concept of a ‘balanced diet’ (INL) | 0 | 0 | 0 | 1 | 1 |
|  | I use the internet when I am looking for information about nutrition such as diet (FNL). | 0 | 0 | 0 | 1 | 1 |
|  | I discuss diet with my friends, family, and relatives (INL). | 0 | 0 | 0 | 1 | 1 |
|  | I have changed my eating habits based on the information about diet that I have gathered (CNL). | 0 | 0 | 0 | 1 | 1 |
|  | I often read material about what constitutes a balanced diet (FNL). | 0 | 0 | 0 | 1 | 1 |

|  |  |  |  |  |  |
| --- | --- | --- | --- | --- | --- |
| I would readily get involved in issues targeted at improving people's diet in my community in COUNTRY (CNL). | 0 | 0 | 0 | 1 | 1 |
| I am willing to take an active role in measures aimed at promoting a healthier diet at my school/college/workplace/community (CNL). | 0 | 0 | 0 | 1 | 1 |
| I expect my school/college/workplace to serve healthy food (INL). | 0 | 0 | 0 | 1 | 1 |
| I try to influence others (for example my family and friends) to eat healthy food (INL). | 0 | 0 | 0 | 1 | 1 |

## Declarations

### Ethical approval and consent to participate

This study was conducted in accordance with the 1964 Declaration of Helsinki and ethical approval was granted by the Research and Ethics Committee at the School of Public Health, Makerere University (Reference number: SPH-2023-460). Written informed consent was obtained from parents/guardians of adolescents (10-17 years), followed by adolescent assent. Adults (18+ years) provided written informed consent.

### Consent for publication

Not applicable.

### Authors’ contributions

Conceptualization: TB, EB, JB, RN and DG; Data curation: TB; Formal analysis: TB; Funding acquisition: DG; Investigation: TB; Methodology: TB, EB, JB, MM, JK, RN, and DG; Validation: TB, EB, JB, RN and DG; Visualization: TB, EB, JB, MM, JK, RN and DG; Writing original draft: TB; Writing, review and editing: All authors reviewed and edited the published version of the manuscript.

## Acknowledgements

The authors would like to thank the adolescents and young adults who participated in the study and the community health workers who assisted as community guides during data collection.

